# Epidemiological and cohort study finds no association between COVID-19 and Guillain-Barré syndrome

**DOI:** 10.1101/2020.07.24.20161471

**Authors:** S Keddie, J Pakpoor, C Mousele, M Pipis, PM Machado, M Foster, CJ Record, YS Keh, J Fehmi, RW Paterson, V Bharambe, LM Clayton, C Allen, O Price, J Wall, A Kiss-Csenki, DP Rathnasabapathi, R Geraldes, T Yermakova, J King-Robson, M Zosmer, S Rajakulendran, R Nortley, CR Marshall, E Newman, N Nirmalananthan, G Kumar, AA Pinto, J Holt, TM Lavin, K Brennan, M Zandi, DL Jayaseelan, J Pritchard, RDM Hadden, H Manji, HJ Willison, S Rinaldi, AS Carr, MP Lunn

## Abstract

**Background:** Reports of Guillain-Barré Syndrome (GBS) have emerged during the Coronavirus Disease 2019 (COVID-19) pandemic. This epidemiological and cohort study sought to investigate any causative association between COVID-19 infection and GBS.

**Methods:** The epidemiology of GBS cases reported via the UK National Immunoglobulin Database were studied from 2016-2019 and compared to cases reported during the COVID-19 pandemic. For the cohort study, members of the British Peripheral Nerve Society reported all cases of GBS during the pandemic. The clinical features, investigation findings and outcomes of COVID-19 (definite or probable) and non-COVID-19 associated GBS cases were compared.

**Results:** The UK GBS incidence from 2016-2019 was 1.65-1.88 per 100,000 people per year. GBS and COVID-19 incidence varied between regions and did not correlate (r = 0.06, 95% CI −0.56 to 0.63, p=0.86). GBS incidence fell between March and May 2020 compared to the same months of 2016-2019. Forty-seven GBS cases were included in the cohort study (13 definite, 12 probable COVID-19 and 22 non-COVID-19). There were no significant differences in the pattern of weakness, time to nadir, neurophysiology, CSF findings or outcome. Intubation was more frequent in the COVID-19+ve cohort (7/13, 54% vs 5/22, 23% in COVID negative) thought to be related directly to COVID-19 pulmonary involvement.

**Conclusions:** This study finds no epidemiological or phenotypic clues of SARS-CoV-2 being causative of GBS. GBS incidence has fallen during the pandemic which may be the influence of lockdown measures reducing transmission of GBS inducing pathogens such as *Campylobacter jejuni* and respiratory viruses.

## Introduction

The first cases of severe acute respiratory syndrome coronavirus 2 (SARS-CoV-2) were reported to the World Health Organisation in late 2019, and by March 2020 COVID-19 was pandemic.^1^ SARS-CoV and Middle East respiratory syndrome coronavirus (MERS-CoV) were associated with neurological sequelae. ^2^ Early reports identified neurological symptoms of COVID-19 infection as headache, anosmia and dysgeusia.^3^ Subsequently COVID-19 infection has been associated with stroke, meningoencephalitis, acute disseminated encephalomyelitis and Guillain-Barré syndrome (GBS).^4–6^ The first reported case series of five patients with GBS following SARS-CoV-2 infection were reported in April 2020.^7^ We have identified 18 further reports of GBS associated with COVID-19 (see supplementary material).

GBS is an acute, post-infectious immune mediated polyradiculoneuropathy typically arising a few days to 6 weeks after bacterial or viral infections including influenza, *Campylobacter jejuni, Haemophilus influenzae*, Mycoplasma pneumoniae, Epstein-Barr virus, cytomegalovirus, and more recently, Zika virus.^8–10^ The pathological initiating event of GBS is a humoral response, where antibodies to surface epitopes of the invading pathogen cross-react with peripheral nerve glycolipids as a result of molecular mimicry, resulting in complement fixation, macrophage attraction and resultant peripheral axon or myelin nerve damage. ^11^ This mechanism is definitively established for *C. jejuni* associated GBS,^12,13^ and with less certainty for other pathogens. SARS CoV-2 is a single-stranded RNA enveloped virus. Open reading frames (ORF) encode for replicase proteins and the structural proteins which are the spike (S), nucleocapsid (N), envelope (E) and membrane (M) proteins. ^14^ To date we know of no homology between SARS-CoV-2 surface epitopes and peripheral nerve tissue. Reports of diverse anti-ganglioside antibodies suggest a uniform immune mediated hypothesis is unsupported. More comprehensive epidemiological characterisation is crucial to understanding any causal link. With over 13 million cases of COVID-19 infection worldwide by 19^th^ July,^15^ the question of whether COVID-19 infection is a cause of GBS or a coincidental finding remains to be answered.

We performed a UK population-based epidemiological study to investigate any causal relationship between COVID-19 infection and GBS. We furthermore characterised a cohort of UK cases of COVID-19 associated GBS to explore any difference from non-COVID-19 GBS. Finally, we explored any homology between SARS-CoV-2 and the human genome and proteome which would support the molecular mimicry mechanism.

## Methods

### Epidemiological case reporting

Cases of GBS were ascertained from the National Immunoglobulin Database from the 1^st^ January to the 31^st^ May 2020. NHS England (NHSE) procures the total intravenous immunoglobulin (IVIg) supply for England, Scotland and Northern Ireland. NHSE mandates every IVIg prescription is approved by a clinical panel and reported onto the database within 90 days. Recording compliance is almost 100% as hospital trusts are only reimbursed once records of dispensed volumes are submitted; these are retrospectively cross-checked against supply and returned stocks. ^17^ Current UK guidance for GBS treatment indicates IVIg and plasma exchange (PLEX) as first line therapy, ^18^ but IVIg is in practice first line in most UK hospitals due to lack of PLEX resources. The International GBS Outcome Study (IGOS) reported 1000 GBS cases, within which 86% (612/715) of European cases were treated with IVIg, 6% PLEX and 7% received no treatment.^19^ In reported COVID-19 GBS cases (supplementary material), 97% (34/36) received IVIg. To ensure complete reporting at the inception date, NHSE specifically mandated all users of the National Immunoglobulin database by email to log any outstanding GBS cases by 30^th^ June 2020. Data retrieval was performed on the 7^th^ July 2020 to allow time for reporting delay.

We searched the NHSE IVIg database for GBS cases from 2016 to 2019 to determine the incidence of non-COVID-19 reported cases of GBS to compare to the 2020 pandemic data. Data were stratified by hospital trust and region. UK population data for COVID-19 infection were collated from Public Health England, Health Protection Scotland and the Public Health Agency of Northern Ireland.

### Cohort study

In parallel, we conducted a cohort study to compare the phenotype of COVID-19 associated GBS (definite and probable) to COVID-19 negative GBS reported during the same study period.

Reports of GBS were submitted by members of the British Peripheral Nerve Society, who cover 81 different UK sites. Members were emailed on a weekly basis to collect information on hospital presentations of GBS from the 1st March to the 31^st^ May, 2020. Reporting was restricted to BPNS members to achieve a comprehensively characterised representative sample of incident cases diagnosed by peripheral nerve experts. Data were entered to the International Neuromuscular COVID-19 database (www.ucl.ac.uk/centre-for-neuromuscular-diseases/news/2020/may/international-neuromuscular-covid-19-database), at the Centre for Neuromuscular Disease. Cohort study data collection ended on July 1^st^ 2020 to allow time for retrospective case reporting. Anonymised clinical data of demographics and medical history, COVID-19 infection, symptoms and management were collected. Precipitating illness, clinical features of GBS, investigation findings including cerebrospinal fluid (CSF) and electrophysiology, management and outcomes were also collated.

To cross validate data collected from the cohort study, the phenotypic characteristics were compared against the published International GBS Outcome Study cases to see whether pandemic presentations differed from non-pandemic phenotypes.^19^

### Evidence of COVID-19

GBS cases were stratified into three groups; definite COVID-19, probable COVID-19 and non-COVID-19. Definite cases had either a positive nasal or throat swab PCR for viral RNA or a subsequent positive serological test for anti-SARS-CoV-2 IgM or IgG irrespective of clinical signs and symptoms. Probable cases were defined by the presence of clinical symptoms consistent with COVID-19 infection as per the European Centre for Disease Prevention and Control case definitions, ^20^ or pulmonary imaging (CXR or CT) highly suggestive of COVID-19 (airway opacification typically bilateral, peripheral and basal in distribution) where PCR analysis was negative. An association of COVID-19 with GBS was defined by an onset of GBS within 8 weeks of acute infection (clinically or on confirmed laboratory findings).

### Search for homology between SARS CoV-2 and human genome and proteome

We searched for evidence of molecular mimicry between SARS CoV2 proteins and human nerve axonal or myelin proteins and glycoproteins. We searched for human homologs of proteins encoded by the SARS CoV-2 genome using the National Centre for Biotechnology Information (NCBI’s) Basic Local Alignment Search Tool (BLAST) to identify common amino acid sequences in the human Reference Sequence Database (refseq_protein). The NCBI BLAST was also used to query the SARS CoV-2 genome against the human genome for any significant alignments at specific genomic loci.

The expect value (E-value) quantifies the number of times a specific alignment can be ‘expected’ to occur in a database by chance. As the E-value decreases the significance of the alignment in the specified database increases. Any alignment with an E-value of < 1×10^−4^ was considered homologous to a human protein (error rate < 0.01%).

### Statistical analysis

The incidence rates of GBS and COVID-19 (95% confidence intervals by Byar’s approximation method^22^) were calculated by dividing regional cases and time period by the relevant mid-year population estimate. Mid-year population estimates at both regional and national level were obtained from the UK Office for National Statistics. ^21^ We explored any association between the incidence of GBS and the incidence of COVID-19 in UK regions in 2020 (January to May) using Pearson’s correlation coefficient. The Shapiro-Wilk test was used to determine suitability of parametric tests. COVID-19 definite and probable cases were compared against non-COVID-19 associated GBS for statistical analysis. Secondary analysis was performed to compare data from the entire cohort against the IGOS study cohort to determine whether characteristics differed between pandemic and non-pandemic GBS phenotypes. We used one Mann-Whitney U to test non-parametric continuous data, and the χ^2^ or Fisher’s exact test to compare proportions. R (4.0.0) and GraphPad prism (8.1.2) were used for analysis and figures.

### Ethics

The UK Health Research Authority was consulted and advised the study did not require review by an NHS Research Ethics Committee as an analysis of previously collected non-identifiable information. The project was submitted as a “Service Evaluation” to the Clinical Audit and Quality Improvement Subcommittee (CAQISC).

## Results

### Epidemiological study

The NHSE IVIg Database reported a mean of 1098 (range 1021-1155) GBS cases per year between 2016-2019 (monthly range 83 to 170 cases). Annual UK GBS incidence was 1.65-1.88 per 100,000 people each year across this period, consistent with the incidence of GBS in Europe and North America from a meta-analysis of 1,643 GBS cases (range 0.81 to 1.89 cases per 100,000). ^23^ These figures along with the mandatory reporting supports the database as a complete and accurate resource for epidemiological analysis.

Although COVID-19 was first reported in the UK on January 31^st^ 2020, significant numbers of daily new infections (>1000 per day) did not occur until March, with the highest recorded daily count of 6,201 confirmed cases on 1st May. ^24^ Through April and May there were 4000-6000 COVID-19 cases per day. As a result, COVID-19 GBS cases were expected to rise in subsequent weeks if a strong association existed (see Figure 1). However, even accounting for the consistent summer dip in GBS cases, GBS cases in March (93), April (70) and May (56) were significantly fewer than previous years (mean 132 (March), 116 (April) and 113 (May) from 2016 to 2019) (Figure 2). GBS and COVID-19 incidences varied across the UK with no correlation at a regional level (r= 0.060 95% CI −0.56 to 0.63, P=0.86) (See Figure 3 and supplementary table 2).

**Figure 1:**
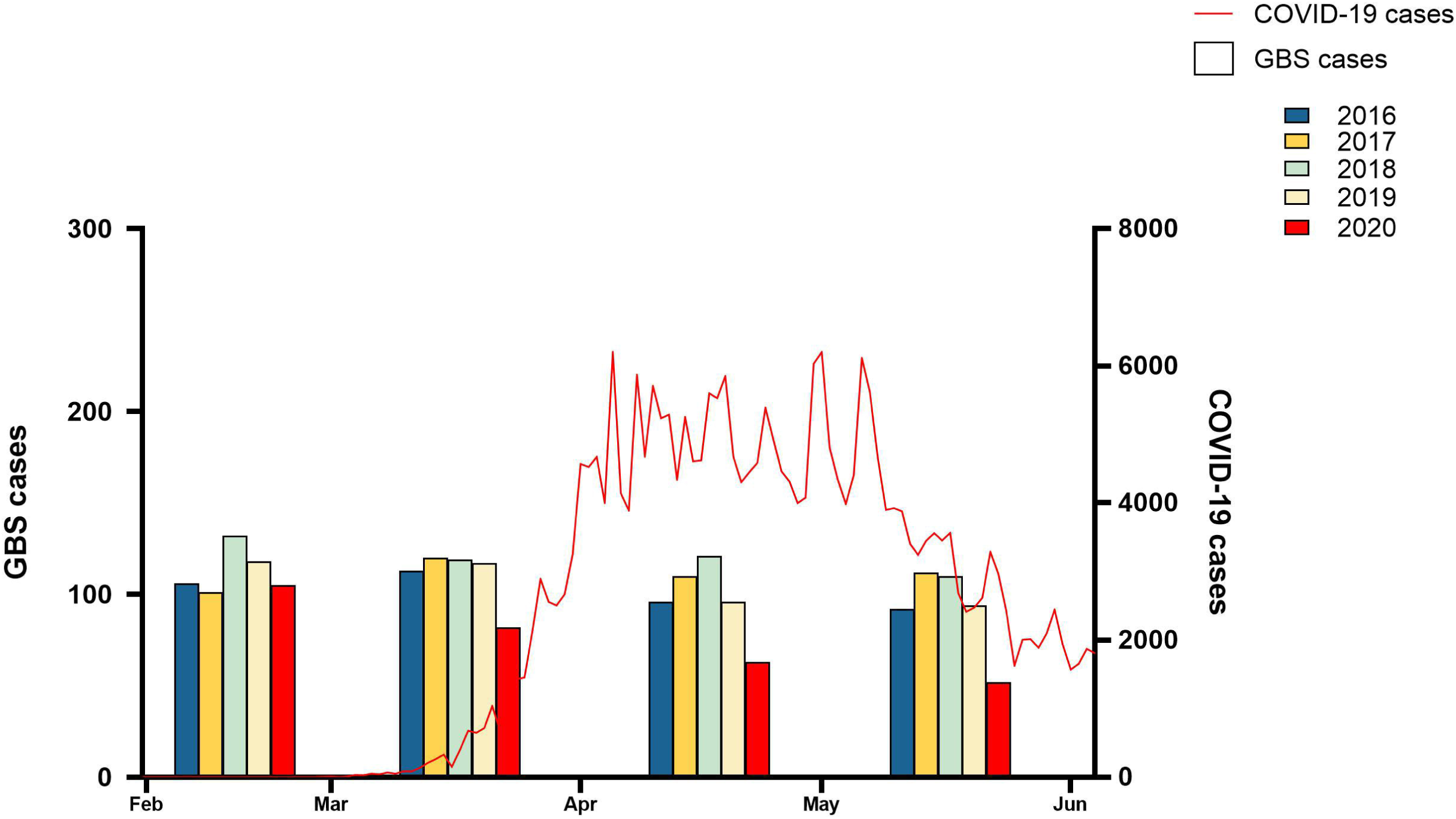
Numbers of new daily COVID-19 infections from February to June 2020 (red line) compared to Guillain-Barré syndrome cases in the UK between February and June from 2016 to 2020 (years depicted by colours in figure legend).

**Figure 2:**
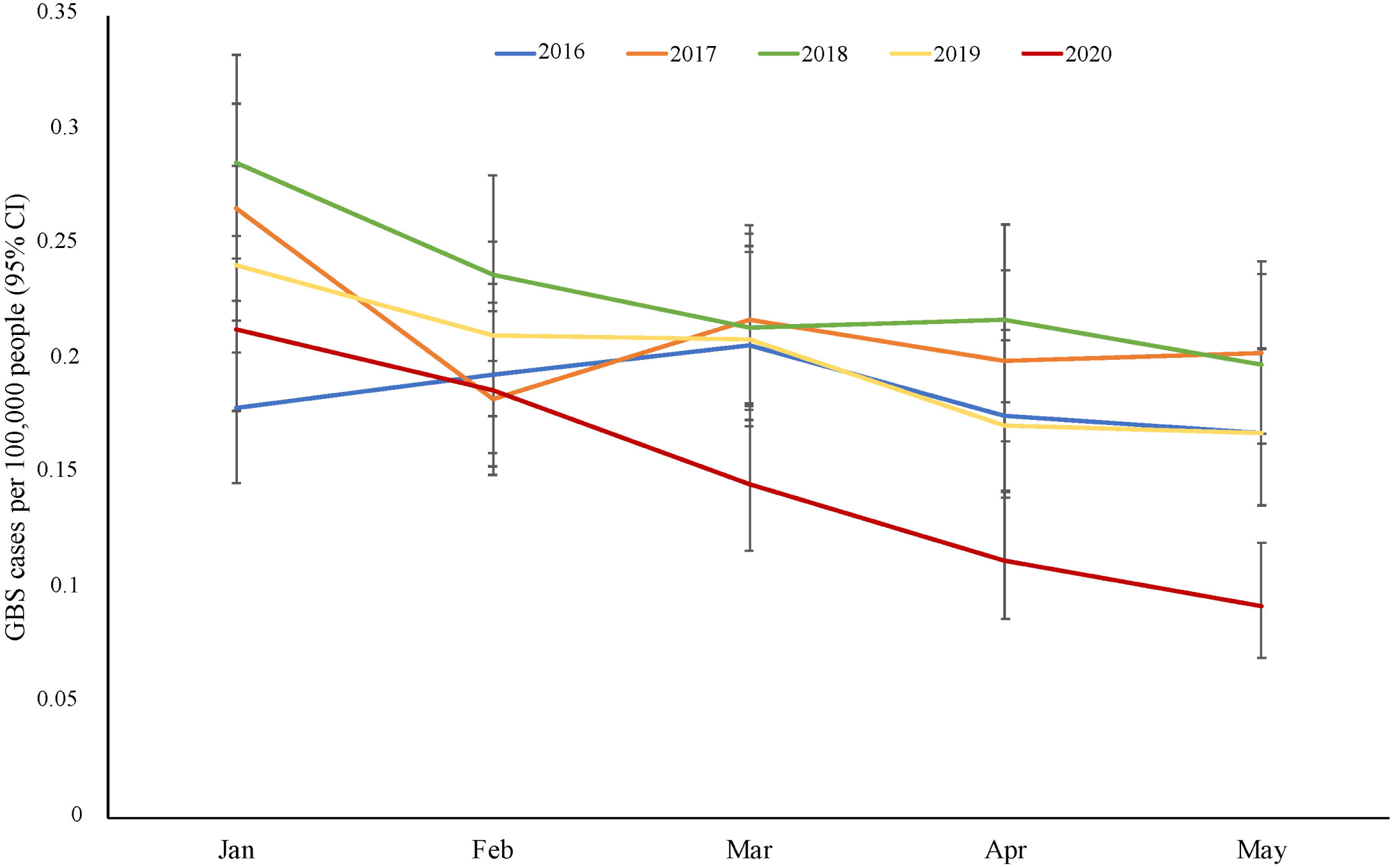
Monthly incidence of Guillain-Barré syndrome per 100,000 people treated with IVIg in the UK between January and June for 2016-2020.

**Figure 3:**
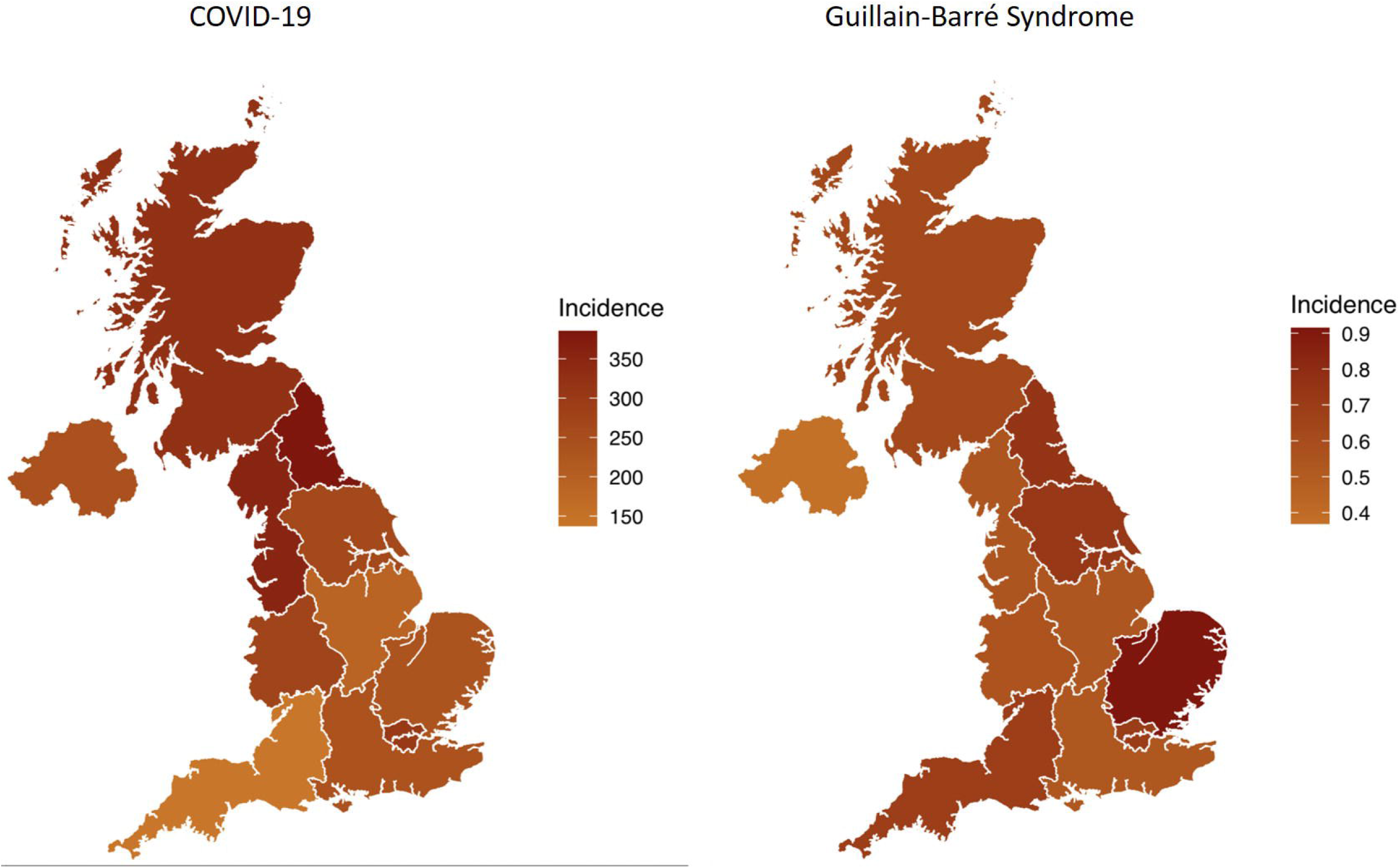
Heatmap of regional incidences of Guillain-Barré syndrome and COVID-19 infections per 100,000 across the UK from January to June 2020.

### Cohort study

Forty-seven cases of GBS were reported to the cohort. Of these,13 had definite COVID-19 infection, 12 were probable, and 22 had GBS with no evidence of COVID-19. Median age was 57 years (IQR 19-88), 33 were men (70%) and 29 (66%) were Caucasian. The male:female ratio in the COVID-19 patients was 5.5 compared to 1.4 in the non-COVID and IGOS study. Men are more likely to be significantly unwell and hospitalised with COVID-19 infection which may partially or completely explain this difference.^25,26^

The clinical characteristics of the patients with GBS are shown in Table 1. Of the patients with COVID-19 infection, the median time between onset of infective symptoms and neurological weakness was 12 days for definite and 5 for probable cases; however the range of time intervals was very broad, ranging from 0 to 37 days in definite and −14 to 52 days in probable cases. Three probable cases developed symptoms of GBS without any clear COVID-19 symptoms, and had incidental imaging evidence of COVID-19 suggesting recent mild or asymptomatic infection. Numbers of non-COVID-19 GBS cases with symptoms of a precipitating illnesses, particularly gastroenteritis, were significantly fewer than that compared to the IGOS cohort (1/22, 5% in non-COVID-19 cases compared to 229/857, 27% in the IGOS cohort, p=0.014) This may be an effect of lock down with improved hand hygiene reducing numbers of faecal-oral pathogen transmissions. The pattern of weakness and time to nadir were no different between COVID-19 associated GBS and non-COVID-19 GBS. Cranial nerve involvement was the only finding more frequent in the IGOS study compared to this cohort, possibly due to differences in data acquisition of the two reporting systems. Although not statistically significant, electrophysiological determination revealed a higher proportion of axonal GBS in the non-COVID-19 patients (four Acute Motor and Sensory Axonal Neuropathy (AMSAN) and one Acute Motor Axonal Neuropathy (AMAN), 23%), compared to one with AMSAN from the COVID-19 positive group.

**Table 1:** Cohort study data demonstrating clinical characteristics of Guillain-Barré Syndrome with definite, probable and without COVID-19 infection.

| GBS features | Overall | COVID-19<br>Definite | COVID-19<br>Probable | Non<br>COVID-19 | IGOS cohort<br>2018 | P Value |
| --- | --- | --- | --- | --- | --- | --- |
| **Number** | 47 | 13/47 (28) | 12/47 (26) | 22/47 (47) | 925 |  |
| **Age, years (IQR)** | 58 (57 - 66) | 60 (57 - 66) | 57 (50 - 60) | 54.5 (34-66) | 51 (33-64) | 0.528 |
| **Sex, male:female (ratio)** | 33:14 (2.36) | 11:2 (5.5) | 9:3 (3.0) | 13:9 (1.4) | 552:373 (1.4) | 0.200 |
| **Ethnicity/ Origin (%)** |  |  |  |  |  |  |
| White | 29 (62) | 8 (62) | 6 (50) | 15 (68) |  | 0.548 |
| Black, Asian and minority ethnic | 12 (26) | 3 (23) | 2 (17) | 5 (23) |  |  |
| Unknown | 6 (13) | 2 (15) | 2 (17) | 2 (9) |  |  |
| **Comorbidities (%)** |  |  |  |  |  |  |
| Hypertension | 8 (17) | 3 (23) | 1 (8) | 4 (18) |  | >0.999 |
| Diabetes | 5 (10) | 1 (8) | 1 (8) | 3 (14) |  | >0.999 |
| Hypercholesterolaemia | 6 (13) | 2 (15) | 3 (25) | 1 (5) |  | 0.194 |
| Cerebrovascular Disease | 2 (4) | 1 (8) | 0 (0) | 1 (5) |  | >0.999 |
| COPD/ Asthma | 2 (4) | 2 (15) | 0 (0) | 0 (0) |  | 0.491 |
| **Severity and Distribution of weakness (%)** |  |  |  |  |  |  |
| Tetraparesis | 30 (64) | 9 (69) | 8 (67) | 13 (59) | 677/924 (73) | 0.558 |
| Weakness lower limbs only | 10 (21) | 2 (15) | 3 (25) | 5 (23) | 105/924 (11) | >0.999 |
| Weakness upper limbs only | 0 (0) | 0 (0) | 0 (0) | 0 (0) | 19/924 (2) | >0.999 |
| Unilateral limb weakness | 1 (2) | 1 (8) | 0 (0) | 0 (0) | 10/924 (1) | >0.999 |
| No limb weakness | 4 (9) | 1 (8) | 1 (8) | 2 (9) | 15/924 (2) | >0.999 |
| Other | 2 (4) | 0 (0) | 0 (0) | 2 (9) |  | 0.213 |
| **GBS syndrome (%)** |  |  |  |  |  |  |
| AIDP | 21 (45) | 7 (54) | 2(17) | 12 (55) | 312/573 (55) | 0.248 |
| Axonal (AMAN/ AMSAN) | 6 (12) | 0 (0) | 1 (8) | 5 (23) | 33/573 (6) | 0.084 |
| Miller Fisher | 1 (2) | 1 (8) | 0 (0) | 0 (0) |  | >0.999 |
| Neurophysiology not assessed | 17 (36) | 5 (38) | 8 (67) | 4 (18) |  | **0.031** |
| Normal Neurophysiology | 2 (4) | 0 (0) | 1 (8) | 1 (5) | 36/573 (6) | 0.084 |
| **Preceding illness (%)** |  |  |  |  |  |  |
| Upper respiratory tract infection | 18 (38) | 10 (77) | 2(17) | 6 (27) | 303/857 (35) | 0.229 |
| Gastrointestinal infection | 1 (2) | 0 (0) | 0 (0) | 1 (5) | **229/857 (27)\*** | 0.468 |
| Other | 2 (4) | 0 (0) | 0 (0) | 2 (9) |  | 0.214 |
| **Presence of sensory deficit (%)** | 24 (51) | 6 (46) | 7 (58) | 11 (50) | 543/890 (59) | >0.999 |
| **Cranial nerve involvement (%)** | 12 (26) | 3 (23) | 6 (50) | 6 (27) | **464/922 (50)\*** | 0.550 |
| **CSF results** |  |  |  |  |  |  |
| **Protein, g/l (IQR)** | 0.70 (0.47–1.23) | 0.78 (0.50–0.99) | 1.08 (0.69-1.66) | 0.49 (0.33-1.23) |  | 0.172 |
| <0.4 g/l | 9/40 (23) | 0/11 (0) | 2/10 (20) | 7/19 (37) |  |  |
| 0.4-1 g/l | 17/40 (42) | 8/11 (73) | 3/10 (30) | 6/19 (32) |  |  |
| >1 g/l | 14/40 (36) | 3/11 (27) | 5/10 (50) | 6/19 (32) |  |  |
| **White Cell Count, cells/ul (IQR)** | 2 (1 - 2) | 2 (0 - 5) | 2 (0 - 13) | 2 (0 - 38) |  | 0.762 |
| <5 | 36/40 (90) | 10/10 (100) | 8/10 (80) | 16/19 (84) |  |  |
| 5 to 10 | 1/40 (3) | 0/10 (0) | 0/10 (0) | 2/19 (11) |  |  |
| >10 | 3/40 (7) | 0/10 (0) | 2/10 (20) | 1/19 (5) |  |  |
| **Positive Gangliosides (%)** | 5/47 (11) | 0 (0) | 0 (0) | 5 (10) |  | **0.017** |
| **Time from COVID-19 symptoms to weakness, days (IQR)** | NA | 12 (4 - 21) | 5 (-7 - 19) | NA |  |  |
| **Time weakness to admission, days (IQR)** | 4 (1 - 8) | 2 (-1 - 4) | 10 (4 - 14) | 4 (1 - 5) | 3 (2 - 6) | 0.534 |
| **Time from weakness onset to nadir, days (IQR)** | 10 (5 - 20) | 7 (4 - 11) | 11 (5 - 17) | 6 (4 - 9) |  | 0.311 |
| **Invasive mechanical ventilation during illness (%)** | 12 (26) | 7 (54) | 0 (0) | 5 (23) | 176/925 (19) | 0.747 |
| **ITU as the maximum level of care required (%)** | 17 (36) | 7 (54) | 1 (8) | 9 (41) |  | 0.558 |
| **HDU as the maximum level of care required (%)** | 4 (9) | 3 (23) | 0 (0) | 1 (5) |  | 0.611 |
| **GBS disability score at 4 weeks from admission, (IQR)** | 3 (2 - 4) | 3 (2-4) | 2.5 (1.75 - 3) | 3.5 (2 - 4) |  | 0.990 |
| Healthy - 0 | 3 (6) | 0 (0) | 2 (17) | 1 (5) |  |  |
| Minor symptoms and capable of running - 1 | 4 (9) | 1 (8) | 1 (8) | 2 (9) |  |  |
| Able to walk 10 m without assistance but unable to run - 2 | 9 (19) | 2 (15) | 3 (25) | 4 (18) |  |  |
| Able to walk 10 m across an open space with help - 3 | 10 (21) | 3 (23) | 4 (33) | 3 (14) |  |  |
| Bedridden or chair bound - 4 | 14 (30) | 4 (31) | 2 (17) | 8 (36) |  |  |
| Requiring assisted ventilation for at least part of the day - 5 | 4 (9) | 1 (8) | 0 (0) | 3 (14) |  |  |
| Dead - 6 | 1 (2) | 1 (8) | 0 (0) | 0 (0) |  |  |
| **GBS disability score on discharge, (IQR)** | 3 (2 - 4) | 3 (2 - 4) | 2 (1.75 - 3) | 3 (2 - 4) |  | 0.658 |
| Healthy - 0 | 3 (6) | 1 (8) | 1 (8) | 1 (5) |  |  |
| Minor symptoms and capable of running - 1 | 4 (9) | 1 (8) | 1 (8) | 2 (9) |  |  |
| Able to walk 10 m without assistance but unable to run - 2 | 11 (23) | 2 (15) | 4 (33) | 5 (23) |  |  |
| Able to walk 10 m across an open space with help - 3 | 10 (21) | 3 (23) | 3 (25) | 4 (18) |  |  |
| Bedridden or chair bound - 4 | 8 (17) | 2 (15) | 1 (8) | 4 (18) |  |  |
| Requiring assisted ventilation for at least part of the day - 5 | 1 (2) | 1 (8) | 0 (0) | 0 (0) |  |  |
| Dead - 6 | 1 (2) | 1 (8) | 0 (0) | 0 (0) |  |  |
| **Initial Treatment Type (%)** |  |  |  |  |  |  |
| IVIg | 39 (83) | 9 (69) | 9 (75) | 21 (95) |  | 0.052 |
| PLEX | 0 (0) | 0 (0) | 0 (0) | 0 (0) |  |  |
| Nil | 8 (17) | 4 (31) | 3 (25) | 1 (5) |  | 0.052 |
| **Mortality (%)** | 1 (2) | 1 (8) | 0 (0) | 0 (0) | 44/659 (7) | >0.999 |

Data are presented as *n* (%) or median (IQR). *P*-values represent a comparison between COVID-19 definite and COVID-19 probable to non-COVID-19 GBS. *P*-values below 0.05 are highlighted in bold. \* Represents a statistically significant difference ( $p < 0.05$ ) between the COVID-19 cohort (definite, probable and non-COVID-19) compared to that of the IGOS study cohort. Note that IGOS denominator values reflect worldwide cases reported ( $n=925$ ) or once stratified for 'Europe/American' cases ( $n=715$ ) if available.

The use of ventilation did not differ significantly between COVID-19 (definite and probable) cases vs non-COVID-19 GBS. However, the proportion of COVID-19 definite GBS cases ventilated was higher than all other groups (7/13, 54%). Despite this, the GBS disability score at four weeks was no different between groups, suggesting the requirement for initial ventilation was secondary to active COVID-19 pulmonary involvement in PCR positive definite cases rather than neuromuscular weakness.

There were no differences in the treatment of GBS subgroups. IVIg was the first therapy in 83% cases, and only one patient received plasma exchange as second line therapy. One patient received more than one course of IVIg. One patient (COVID-19 definite) died in the cohort. Death was attributed to pulmonary complications rather than neuromuscular weakness.

### Comparison between SARS-CoV-2 and human genome and proteome

When we examined the entire SARS CoV-2 genome (29903 bases, b; NC_045512.2) as well as overlapping fragments of 1000b (+ 500b) and compared this to the human genome, we found no significant similarity.

We also explored individual proteins encoded by the SARS-CoV-2 genome comparing these against all referenced human proteins. Only the replicase ORF1ab/ORF1a polyprotein (7096 amino acids) produced a match with the human mono-ADP-ribosyltransferase (PARP14) protein. PARP14 belongs to an enzyme superfamily involved in histone modification during DNA damage and is ubiquitously expressed making it unlikely as a mimotope. These two proteins are 32% identical (E-value 3×10^−6^) but have only 1 contiguous identical sequence of 5 or more amino acids (Val-Val-Val-Asn-Ala). The remaining SARS-CoV-2 proteins including the spike/surface, envelope, membrane and nucleocapsid phosphoprotein have no significant similarity with any referenced human protein.

## Discussion

This epidemiological and cohort study does not support any causal link between COVID-19 infection and GBS.^16^ The population based data finds no plausible temporal relationship between COVID-19 and GBS (figure 1), a reduction in cases of GBS in comparison to preceding years (figure 2) and no correlation between COVID and GBS incidence at regional level (figure 3). In addition, there are no scientific data to support a molecular mimicry link of SARS-CoV-2 to GBS at the nucleic acid or protein level, other than a presumptive analogy to other known bacterial and viral GBS-causing pathogens. The lack of even a short, linear homology between the SARS-CoV-2 structure proteins and any axonal or myelin surface proteins would indicate that molecular mimicry with SARS-CoV-2 is unlikely to contribute to the pathogenesis of GBS.

The strong concordance between UK GBS incidence from the NHSE National Immunoglobulin database and reported incidence rates from multinational population based epidemiological studies as well as its mandated collection methods, supports this as an appropriate repository for epidemiological analysis. From this we identified significantly fewer cases of GBS during the COVID-19 pandemic compared to previous years. This could represent an under-ascertainment of cases during lock down for a number of reasons including incomplete IVIg prescription recording. However, UK Hospital Trusts are mandated to report all IVIG treatments to the National Immunoglobulin Database within 90 days. Previous database analyses have shown 95% of cases are recorded within 30 days of GBS treatment, and in 98% within 90 days. We collected data from 1^st^ January to 31^st^ May 2020 on July the 7^th^ allowing sufficient time to capture the majority of reported cases. A subsequent direct check of complete IVIG reporting, specific requests for clinicians to document cases and a cross check between clinical reports and IVIg prescribing data ensured we have as complete a dataset as possible.

Mildly symptomatic patients may have decided not to visit hospital for fear of contracting COVID-19. This issue was recognised in Stroke and Emergency Medicine with declines in overall admissions worldwide. ^27–30^. Unwell frail patients may have not been actively investigated or treated, or not admitted into hospital. Physicians’ prescribing behaviour could also have changed during the pandemic, being more selective in patients treated with IVIg. Even with the above, the National Immunoglobulin Database only records GBS cases meeting criteria for treatment and thus mild GBS phenotypes would not have been reported in 2016-2019, reducing the likelihood of a disparity resulting from mild disease attendance bias. The indication for IVIg treatment in GBS is for non-ambulant patients, and therefore it is unlikely such patients remained at home or would not be admitted to hospital. The cohort study also demonstrated 83% of cases were treated with IVIG, providing cross validation of high treatment rates.

We hypothesise that the lockdown measures introduced to prevent COVID-19 transmission have had secondary effects of reducing other common transmissible infective GBS triggers such as upper respiratory tract infections by social distancing and mask wearing, and gastrointestinal illnesses as fewer people dined out and stricter hand hygiene was implemented. Other studies have reported significant reductions in airborne or faeco-oral transmissible infectious diseases during lockdown supporting this assertion.^31^ Although the true impact of such measures are unknown, avoidance of precipitating infections would conceivably reduce the incidence of GBS, and may explain a reduction of GBS cases during the pandemic. Successful interventions to lower *Campylobacter* contamination of fresh poultry meat previously reduced hospitalisations for GBS by 13%. ^32^ And the numbers of reported infective symptoms preceding GBS, particularly diarrhoea were significantly fewer compared to the IGOS study, consistent with this hypothesis.

True COVID-19 incidence in the UK is known to have been significantly under-reported. Initially there was lack of available testing except in hospital admissions, and only PCR confirmed cases were reported in the published data. UK serological studies of COVID-19 suggest far higher COVID-19 infection rates in the community than PCR confirmed cases.. For example, by 27^th^ April 2020, there were 26,784 confirmed COVID-19 cases reported in London. Serological data from London blood donors on the same date reported the prevalence of prior SARS-CoV-2 infection as 17.5%, ^33^ equivalent to 1,571,850 people having made a serological response. Under reporting of COVID-19 cases poses limitations to true estimates of incidence, resulting in a potential over-reporting of a COVID-19 GBS association.

Whilst at an epidemiological level we found no increase in GBS linked to the COVID-19 epidemic, our data do not exclude the possibility that SARS-Cov-2 might be a driver of GBS in very rare cases, or that a significant reduction in non-COVID-19 GBS could mask a smaller spike of COVID GBS cases. Early series of COVID-19 associated GBS reported specific disease characteristics suggesting differences from typical AIDP.^7^ This larger cohort revealed no differences in the clinical and neurophysiological features, disease severity and outcomes of COVID-19 and non-COVID-19 associated GBS. A larger proportion of COVID-19 PCR positive GBS cases required mechanical intervention compared to all other groups. The similar rate of neurological recovery across all groups suggests ventilation was related to COVID-19 associated pulmonary involvement rather than neuromuscular deficit at nadir.

Guillain-Barré Syndrome is an acute immune mediated inflammatory polyneuropathy which can result in severe disability and death. Epidemiological and laboratory research have revealed a number of infective pathogens that precede autoimmune peripheral nerve damage thought to represent molecular mimicry. Experiential evidence from previous coronaviruses alerted physicians to the possibilities of links between SARS-CoV-2 and neurological diseases, of which large patient cohorts have now been well documented. This epidemiological and cohort study contradicts a growing number of reports postulating causation between SARS-CoV-2 and GBS, and in fact demonstrates a reduction of GBS cases. Although prompt reporting of disease manifestations and potential associations of COVID-19 is important to inform public health decisions, robust scientific assessment to establish causality versus association is essential to evolve our understanding of this novel viral pathogen and its sequelae.

## Data Availability

Anonymised data available at the request of the first or corresponding author

